# Electronic health record (EHR)-detectable phenotypes of statin intolerance: Prevalence and validation in real-world general practice

**DOI:** 10.1101/2025.11.04.25339533

**Authors:** Shagoofa Rakhshanda, Joel Rhee, Siaw-Teng Liaw, Kerry-Anne Rye, Jitendra Jonnagaddala

**Affiliations:** School of Population Health, UNSW Sydney, Kensington, Australia; Discipline of General Practice, School of Clinical Medicine, Kensington, Australia; School of Biomedical Sciences, UNSW Sydney, Kensington, Australia; SREDH Consortium, Sydney, Australia

## Abstract

**Objectives:** To estimate the prevalence of statin intolerance and evaluate the performance of electronic health records (EHR)-detectable electronic phenotyping algorithms among patients prescribed statins for primary prevention of cardiovascular disease.

**Methods:** Using the Electronic Practice Based Research Network (ePBRN) dataset, published phenotyping algorithms were identified and implemented, a reference standard was developed, and algorithm performance was evaluated.

**Results:** Six phenotyping algorithms were evaluated: Minnesota Combined Rule-Based algorithm, Japan statin induced myopathy (SIMs), USA SIMs, Singapore SIMs (A–D), Japan statin-associated muscle toxicity (SAMT), and NHS-UK statin intolerance pathway. Statin intolerance prevalence was 5.09%. No algorithm performed consistently well. Singapore SIMs-B achieved the highest accuracy (57.05%) and sensitivity (92.95%), while Japan SAMT achieved the highest specificity (99.13%) and positive predictive value (76.19%).

**Conclusion:** The estimated prevalence was at the lower end of the reported ranges. Variable algorithm performance highlights the need for further validation before clinical implementation.

## 1. Introduction

Statins or 3-hydroxy-3-methylglutaryl coenzyme A (HMG-CoA) reductase inhibitors are known for their ability to lower cholesterol levels and are used as pharmacological treatments for cardiovascular diseases (CVD). They are used for both primary (preventing the development of CVD) and secondary prevention (preventing the occurrence of further CV events) (1, 2). In Australia, statins are prescribed to 30% of patients aged >55 years and are taken by over 40% of patients aged >65 years (3, 4). In addition to their preventive characteristics, statins are known to cause adverse effects in patients (5). Such adverse effects include muscle damage, and in severe cases, muscle breakdown, which may release myoglobin and lead to renal damage (6). This phenomenon is known as statin intolerance and is characterized by symptoms such as muscle weakness, myopathy (muscle aches, muscle cramps, myositis, and rhabdomyolysis), and increased liver enzymes (7). While statins are generally well tolerated, statin intolerance can occur (either completely or partially) in approximately 5–15% of patients (7, 8, 9). Factors associated with statin intolerance include age, gender, ethnicity, hypothyroidism, obesity, chronic liver disease, diabetes mellitus, and renal failure (5).

At present, there is no established taxonomy to classify statin intolerance; therefore, the condition is defined differently by different scientific groups, trialists, and societies. The International Lipid Expert Panel (ILEP) defines statin intolerance as the inability to tolerate a statin dose required to reduce a patient’s CV risk, which limits the effective treatment of the patient (10). According to the National Lipid Association (NLA), statin intolerance includes any adverse effects on the quality of life that lead to a decrease or cessation of statin use (11). The European Atherosclerosis Society (EAS) defines statin intolerance as the probability of the occurrence of statin-associated muscle symptoms (SAMS), considering the nature of muscle symptoms, elevation in creatine kinase (CK) levels, and their temporal association with statin initiation and discontinuation (12). The Luso-Latin American Consortium (LLAC) and the Canadian Consensus Working Group (CCWG) define statin intolerance as the inability to tolerate two or more statins at any dose or the inability to tolerate increasing doses of statins (13, 14). However, the most recent definition of statin intolerance by the NLA is “one or more adverse effects associated with statin therapy that resolves or improves with dose reduction or discontinuation and can be classified as a complete inability to tolerate any dose of a statin or partial intolerance with inability to tolerate the dose necessary to achieve the patient-specific therapeutic objective. To classify a patient as having statin intolerance, a minimum of two statins should have been attempted, including at least one at the lowest approved daily dosage” (15).

Statin intolerance can be measured or quantified by the number and severity of adverse events/reactions to statin therapy, the severity of reports of muscle pain and weakness, discontinuation of statins due to adverse effects, creatinine kinase values, and symptom measures, including patient-reported myalgia (16, 17). Furthermore, the clinical measures of statin intolerance include the global symptom scale score (Likert scale of symptoms), visual analogue scale (VAS) score (to assess pain), brief pain inventory (BPI), and SAMS-CI scale (18, 19, 20, 21). The management of statin intolerance includes several interventions, including changing the statin dose, switching statins, combining statins with non-statin drug therapy, changing to non-statin drugs, and providing dietary interventions (8, 22, 23).

Identification of statin intolerance can be difficult, not only because of the variability in the definition of the condition but also because of the negative representation of statins by the media, close associates, and other sources (24). Fear of intolerance can lead to nonadherence or discontinuation (5). However, recent studies have shown that this issue can be addressed in electronic health records (EHRs) in primary care using electronic phenotyping (25). Electronic phenotyping involves the systematic identification and categorization of patient health data to create specific disease phenotypic profiles. The process involves using EHR data to classify patients based on the presence or absence of specific health-related traits or diseases by leveraging structured and unstructured data from EHR systems (25, 26). There are two types of electronic phenotyping: rule-based and machine learning-based. Rule-based phenotyping involves using predefined algorithms to classify patients and relies on specified criteria (such as codes and laboratory values) to define a phenotype, which may involve significant manual effort to validate and implement (27, 28). Machine learning-based approaches involve the application of machine learning algorithms to identify phenotypes, with algorithms learning patterns from EHR data to classify different patient groups (25, 29). Rule-based algorithms can be useful for analysing EHR datasets, as the information within these datasets can be implicit or discrepant (30). Thus, to ensure the use of correct information to identify disease cohorts within the EHR system, researchers use rule-based algorithms with defined inclusion and exclusion criteria, which are created in collaboration with clinicians (25, 31). The primary objective of this study was to identify patients with statin intolerance and determine the prevalence of statin intolerance using electronic health records from general practice by implementing rule-based algorithms (or EHR-detectable electronic phenotyping algorithms for statin intolerance) that apply various definitions, criteria, or guidelines for statin intolerance and have been used in different countries. The secondary objective was to validate these algorithms. This study focused on patients who were prescribed statins for the primary prevention of CVD.

## 2. Methods

This study used EHR-detectable electronic phenotyping algorithms for statin intolerance that implement different definitions of statin intolerance to identify patients with statin intolerance

(32). The study methodology was conducted in four stages. First, a comprehensive literature review was undertaken to identify EHR-detectable electronic phenotypes of statin intolerance. Second, these electronic phenotypes were implemented in the dataset of this study. Third, a reference standard was developed through manual review of patient data and subsequently implemented. Finally, the EHR-detectable electronic phenotypes of statin intolerance were validated using established performance measures. The overview of the methodology is presented in **Figure 1**. This study was reported in accordance with the strengthening the reporting of observational studies in epidemiology (STROBE) statement and the reporting of studies conducted using observational routinely collected health data (RECORD) extension for studies using routinely collected health data, with the completed reporting checklists provided in **Supplement 1**.

**Figure 1:**
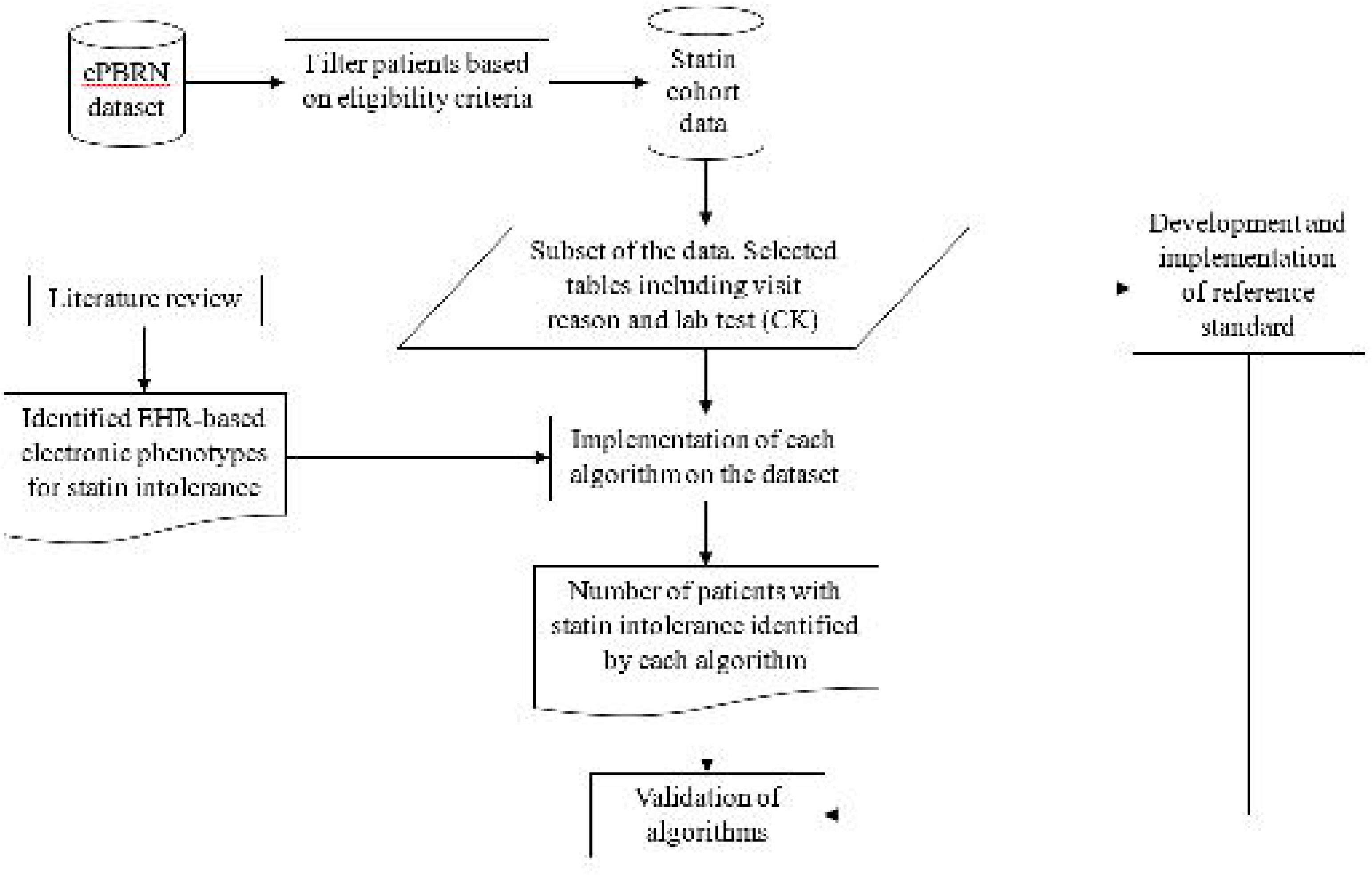
Overview of the study methodology. **Notes:** ePBRN: Electronic Practice Based Research Network; CK: Creatine kinase; EHR: Electronic health record

### 2.1 Dataset

The dataset that was used for this study was the Electronic Practice Based Research Network (ePBRN) Linked Dataset, which is a comprehensive dataset derived from electronic health records (EHRs) from general practice facilities in the Fairfield and Wollondilly regions of the South-Western Sydney local health district (SWS LHD). This dataset contains records for 249,345 patients across 11 general practice sites, who had at least one visit between 2012 and 2019 (33, 34).

The quality of routinely collected data within the ePBRN dataset has been assessed in previous research. An early study assessed the completeness, correctness and consistency of general practice data within the ePBRN dataset. The study found that data quality varied across data domains. Social determinant data were generally well recorded, while greater variability was observed in the recording of risk factors (35). More recently, the quality of smoking information recorded by general practitioners (GPs) within the ePBRN dataset was assessed in terms of plausibility, conformance, and completeness (36). The study showed that routinely collected ePBRN data can be useful for research, while also identifying limitations related to the recording and completeness of some information. Overall, these studies show that the ePBRN is a valuable source of longitudinal real-world primary care data, although the quality and completeness of individual variables need to be considered when using these data for research.

The ePBRN dataset consists of a total of 76 tables categorized according to the GP electronic health record system used (Best Practice or Medical Director). Thirty-nine of the 76 tables are sourced from Best Practice, and 37 tables are sourced from Medical Director. The research objective of this study required the extraction of data on statin intolerance, which is associated with patient diagnosis, procedures, conditions, visit details, and demographics. For this purpose, six tables were strategically chosen from Best Practice (Careplan Goal, Current Rx, Visit Reason, Visits, Past History, and Patients), along with five tables from Medical Director (Diagnosis, Prescription, History, Consultations, and Patients). This selection enhanced the precision and relevance of the experimental data. The extracted data were processed using Python to generate and execute Structured Queried Language (SQL) queries, and a Master table was created to facilitate the application of electronic phenotyping algorithms. The Master table consisted of patients with a recorded statin prescription in the EHR. The prescribing data indicate that a statin was prescribed by a GP but do not confirm whether the medication was subsequently dispensed or taken by the patient, as linked dispensing data were not available for this study.

### 2.2 Study design and population

This was a cross-sectional study using retrospectively collected longitudinal EHR data from 2012 to 2019 in the ePBRN Linked Dataset. The index date was defined as the date of the first recorded statin prescription for each patient. To identify patients with no prior recorded statin use, all available patient records before the index date were searched for evidence of a previous statin prescription. Patients with any recorded statin prescription before the index date were excluded. Rather than applying a fixed lookback period, all available historical records for each patient were used to identify prior statin prescribing and reduce the likelihood of misclassifying previous users as new users. However, as the available history varied between patients, the first recorded statin prescription represents the first prescription record observed in the ePBRN data and may not represent the patient’s first statin use.

Patients were eligible if they were aged 18 years or above at the index date and were observed for at least two years after the index date. Patients were considered active if they had three or more visits for any reason during the observation period of two years following the index date, consistent with the Royal Australian College of General Practitioners (RACGP) definition of active patients (37). This criterion was also applied to ensure that included patients had sufficient ongoing engagement with the general practice to provide adequate longitudinal EHR information for the application of the statin intolerance phenotyping algorithms. Patients with no recorded gender information or birth year, aged over 80 years at the index date, or who did not survive during the observation period were excluded. Moreover, patients who experienced a CVD event before the index date or during the observation period were also excluded. Patients with interacting medicines were identified based on documented adverse events attributed to drug interactions in the available EHR data and were excluded from the study population.

Patients aged over 80 years were excluded because advanced age may introduce clinical complexity and variation in treatment goals, approaches, patterns and outcomes. Moreover, there is limited evidence supporting the benefits of statins for primary prevention of CVD in this age group (38, 39). Patients who did not survive during the observation period were excluded because they did not have the complete two years of observation required for consistent application of the phenotyping algorithms. Patients aged 18 years or older were included because recent reports show that statin therapy is considered for high-risk patients between the ages of 20 and 39 years (40). Patients who experienced a CVD event during the observation period were excluded because the indication for statin therapy changes from primary to secondary prevention following the event, which may alter statin treatment and prescribing patterns. These restrictions were applied to reduce clinical heterogeneity and to focus the study on primary prevention of CVD, where statin prescription and intolerance are more consistent and comparable across individuals.

The observation period for each patient was defined as the two years following the index date. Statin prescription and clinical information recorded during this period were used for the identification of statin intolerance using the electronic phenotyping algorithms.

### 2.3 Identification and implementation of EHR-detectable electronic phenotypes of statin intolerance

A rapid literature review was conducted in PubMed on 18 August 2024 using basic keywords such as “statin intolerance” and “digital definitions” to identify previous studies on EHR-detectable electronic phenotypes of statin intolerance. Relevant publications were screened to identify rule-based algorithms, electronic phenotypes, or clinical pathways that provided operational criteria for identifying statin intolerance and could be implemented using routinely collected EHR data. Inclusion and exclusion criteria for the identified rule-based algorithms were tabulated. The criteria for each algorithm were mapped to the relevant variables available within the ePBRN dataset. The selection of these algorithms was based on their use of structured data elements such as diagnosis codes, medications, and procedure codes for establishing inclusion and exclusion criteria. Approaches that could be mapped to the available ePBRN data were selected for implementation and validation. Statin prescribing information, including statin type, dose, discontinuation, switching, and rechallenge, was identified using prescription records. The CK levels were identified from laboratory results. Muscle symptoms were identified from free text entries in the reasons for visit variable. The codes for the identified phenotyping algorithms were run on the prepared dataset using R version 4.3.2. The number of patients identified by each algorithm was recorded and incorporated into a table. The outcome of interest was statin intolerance status, defined as the identification of a patient as statin intolerant or not statin intolerant according to the criteria of each electronic phenotyping algorithm during the observation period.

### 2.4 Development and implementation of the reference standard

The reference standard was developed using a subset of patients prescribed statins in the ePBRN dataset. This subset consisted of patients who were initially identified as potential cases of statin intolerance by any of the identified electronic phenotyping algorithms (n = 1,369). This approach was used to focus the detailed clinical review on patients with EHR features potentially indicative of statin intolerance and to make manual assessment of the reference standard feasible. Including patients identified by any of the algorithms also allowed potential cases captured by the different phenotypes of statin intolerance to be considered in the reference standard assessment.

Two independent researchers (annotators) manually reviewed the records and classified (annotated) patients according to their statin intolerance status based on the EAS definition of statin intolerance (12). One annotator was a senior researcher with expertise in health informatics, electronic health records, digital disease detection, and primary care research, while the other was a doctoral researcher in population health whose research focused on statin adherence and intolerance using primary care EHR data. The EAS definition was treated as a clinical reference definition rather than an electronic phenotype. The manual review was undertaken to identify reference cases of statin intolerance in accordance with the definition. Any disagreements in classification were discussed and resolved by the annotators.

The EAS definition was selected because it provides a clinically reasoned approach that considers the nature of muscle symptoms and CK elevations (12). The definition recognizes that statin-associated muscle symptoms can occur in the absence of elevated CK. However, for the operational reference standard used in this study, patients with muscle symptoms without elevated CK were not classified as statin intolerant. This more restrictive approach was used to increase the specificity of case identification within routinely collected EHR data, where information on the nature and temporal relationship of muscle symptoms with statin treatment was often incomplete and nocebo/drucebo effects could not be reliably distinguished from pharmacologically mediated symptoms.

Following this, the inter annotator agreement (IAA) between the two researchers was assessed using Cohen’s kappa coefficient. Cohen’s kappa was selected as both annotators independently classified patients using a binary (yes/no) outcome. The Cohen’s kappa coefficient measures the level of agreement between the two annotators while accounting for agreement that may occur by chance. A higher Cohen’s kappa value indicates greater agreement between the annotators (41, 42). Furthermore, the Deviation Score (DS) was also calculated to determine the agreement between the final reference standard and each annotator. The DS for each annotator represented the agreement between that annotator and the reference standard (41, 43). The development and implementation of the reference standard is shown in **Figure 2**. The upper limit of normal (ULN) for CK was defined as 145– 150 U/L for female patients and about 170–250 U/L for male patients. The CK thresholds applied varied according to the criteria of the individual phenotyping algorithms, with some algorithms using CK levels of three times the ULN and others using four times the ULN, depending on the presence or absence of muscle symptoms. The specific CK thresholds applied for each algorithm are presented later in **Table 1**.

**Figure 2:**
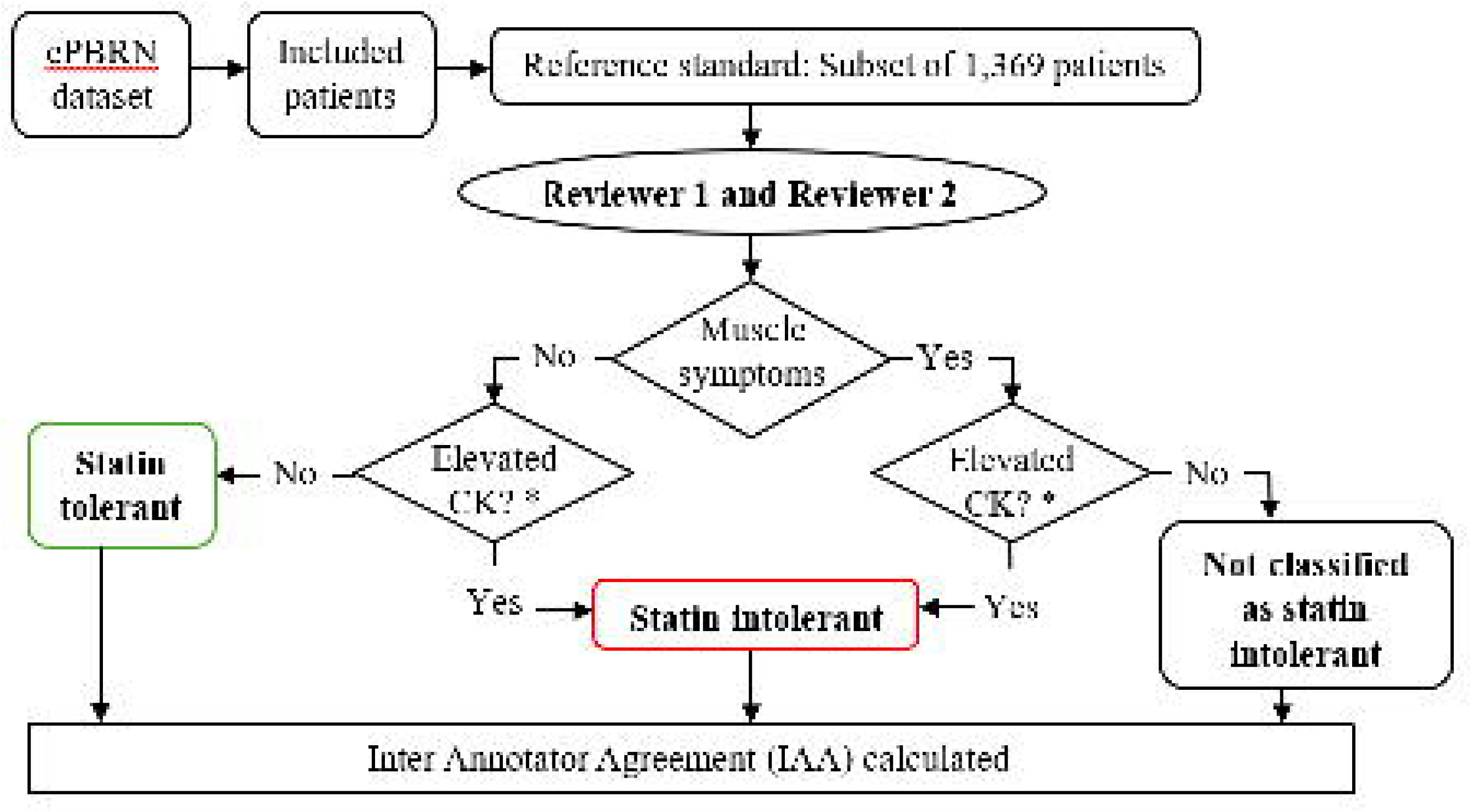
Development and implementation of reference standard. **Notes:** ePBRN: Electronic Practice Based Research Network; CK: Creatine kinase; IAA: Inter Annotator Agreement * The threshold for creatine kinase (CK) was four times the upper limit of normal (ULN) for CK which was defined as 145–150 U/L for female patients and about 170–250 U/L for male patients

**Table 1:**
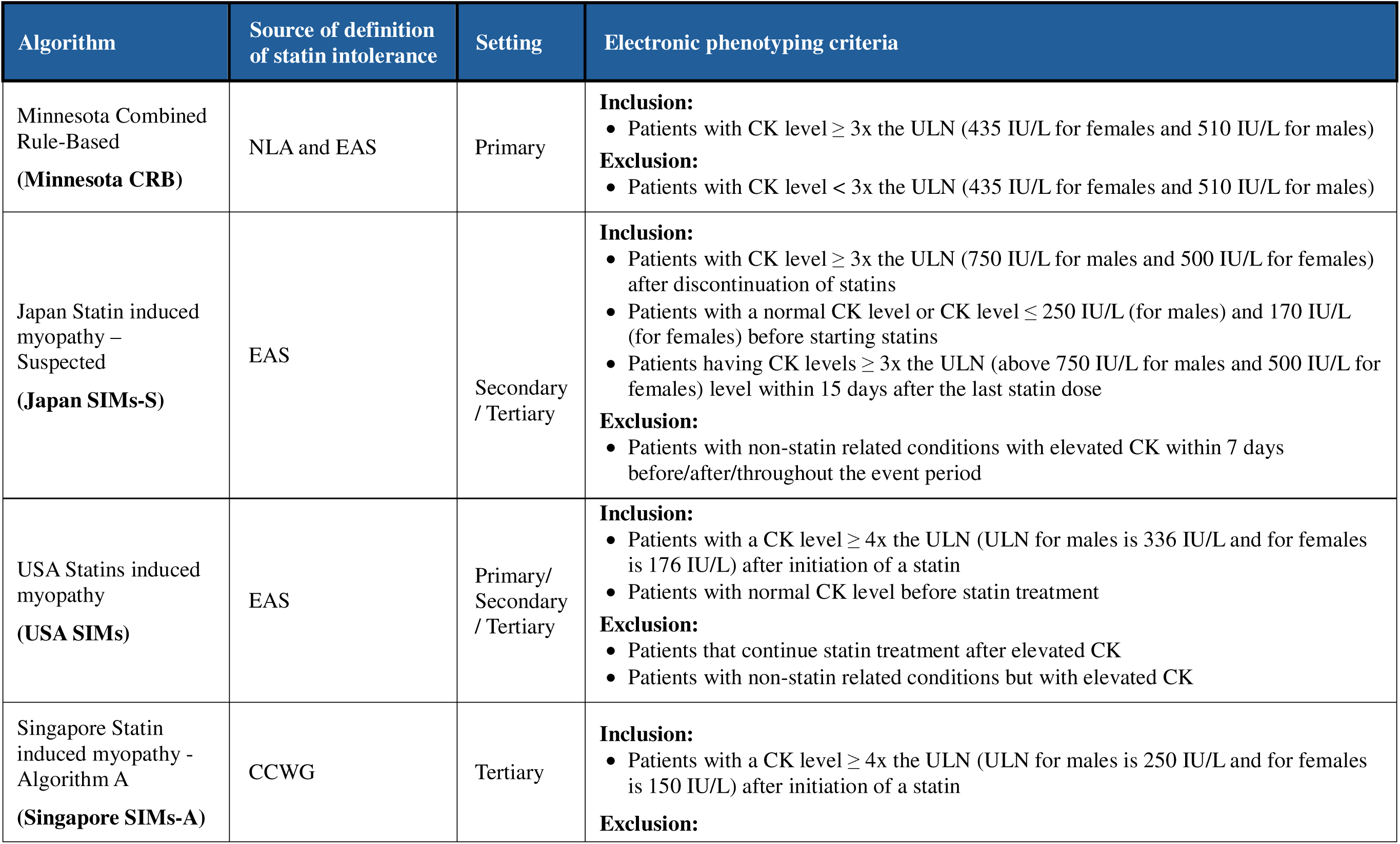
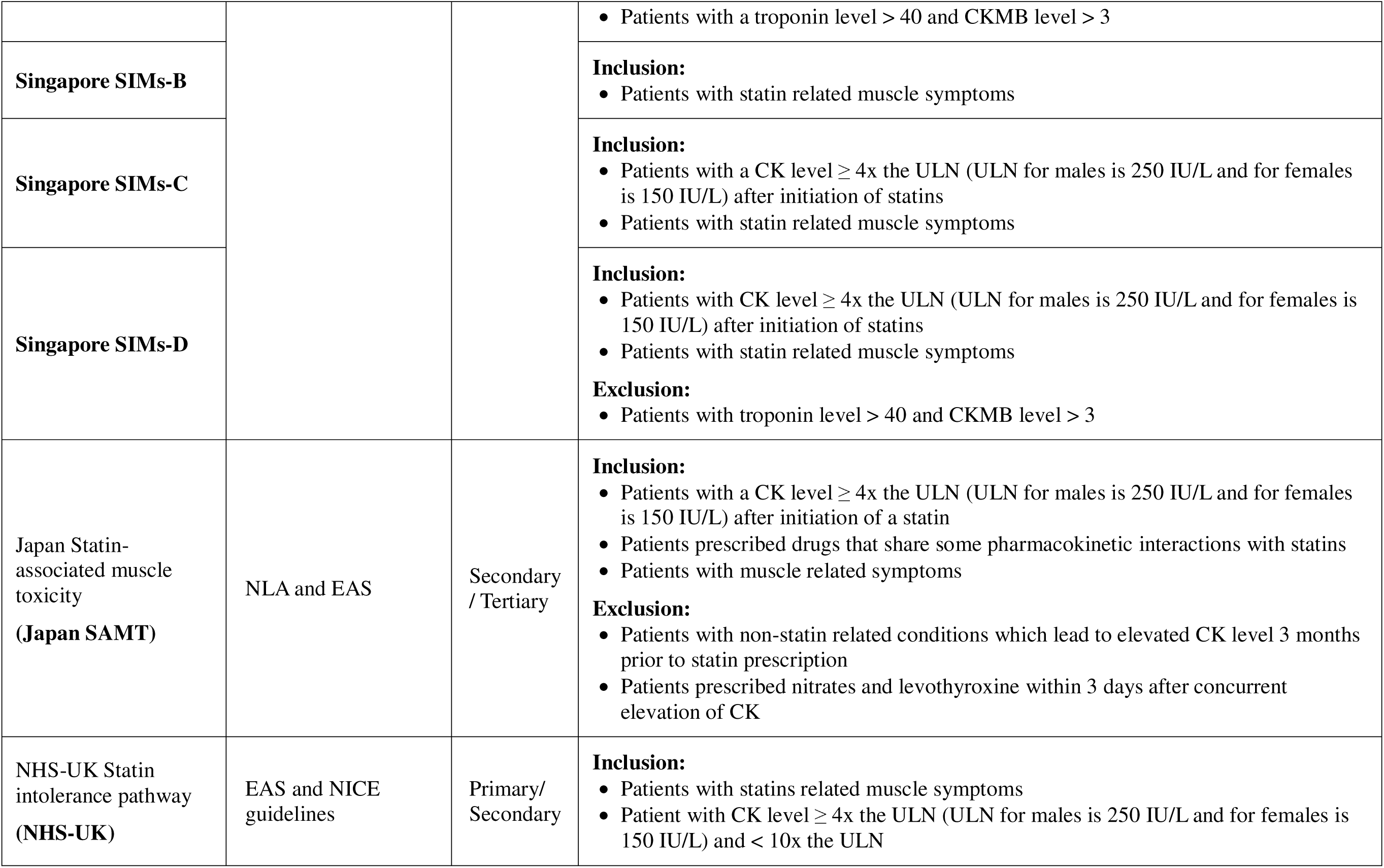
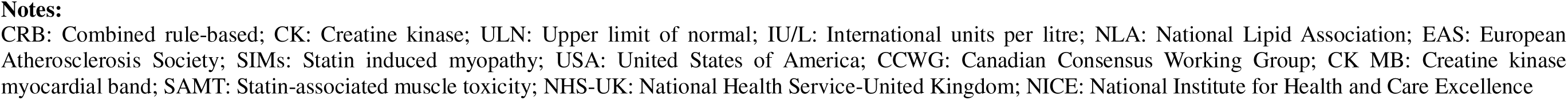
Overview of the identified electronic phenotypic algorithms.

| Algorithm | Source of definition of statin intolerance | Setting | Electronic phenotyping criteria |
| --- | --- | --- | --- |
| Minnesota Combined Rule-Based<br>(Minnesota CRB) | NLA and EAS | Primary | **Inclusion:**<br>Patients with CK level $\geq 3$x the ULN (435 IU/L for females and 510 IU/L for males)<br>**Exclusion:**<br>Patients with CK level $< 3$x the ULN (435 IU/L for females and 510 IU/L for males) |
| Japan Statin induced myopathy – Suspected<br>(Japan SIMs-S) | EAS | Secondary / Tertiary | **Inclusion:**<br>Patients with CK level $\geq 3$x the ULN (750 IU/L for males and 500 IU/L for females) after discontinuation of statins<br>Patients with a normal CK level or CK level $\leq 250$ IU/L (for males) and 170 IU/L (for females) before starting statins<br>Patients having CK levels $\geq 3$x the ULN (above 750 IU/L for males and 500 IU/L for females) level within 15 days after the last statin dose<br>**Exclusion:**<br>Patients with non-statin related conditions with elevated CK within 7 days before/after/throughout the event period |
| USA Statins induced myopathy<br>(USA SIMs) | EAS | Primary/ Secondary / Tertiary | **Inclusion:**<br>Patients with a CK level $\geq 4$x the ULN (ULN for males is 336 IU/L and for females is 176 IU/L) after initiation of a statin<br>Patients with normal CK level before statin treatment<br>**Exclusion:**<br>Patients that continue statin treatment after elevated CK<br>Patients with non-statin related conditions but with elevated CK |
| Singapore Statin induced myopathy - Algorithm A<br>(Singapore SIMs-A) | CCWG | Tertiary | **Inclusion:**<br>Patients with a CK level $\geq 4$x the ULN (ULN for males is 250 IU/L and for females is 150 IU/L) after initiation of a statin<br>**Exclusion:** |
|  |  |  | Patients with a troponin level > 40 and CKMB level > 3 |
| **Singapore SIMs-B** |  |  | **Inclusion:**<br>Patients with statin related muscle symptoms |
| **Singapore SIMs-C** |  |  | **Inclusion:**<br>Patients with a CK level ≥ 4x the ULN (ULN for males is 250 IU/L and for females is 150 IU/L) after initiation of statins<br>Patients with statin related muscle symptoms |
| **Singapore SIMs-D** |  |  | **Inclusion:**<br>Patients with CK level ≥ 4x the ULN (ULN for males is 250 IU/L and for females is 150 IU/L) after initiation of statins<br>Patients with statin related muscle symptoms<br>**Exclusion:**<br>Patients with troponin level > 40 and CKMB level > 3 |
| Japan Statin-associated muscle toxicity<br>**(Japan SAMT)** | NLA and EAS | Secondary / Tertiary | **Inclusion:**<br>Patients with a CK level ≥ 4x the ULN (ULN for males is 250 IU/L and for females is 150 IU/L) after initiation of a statin<br>Patients prescribed drugs that share some pharmacokinetic interactions with statins<br>Patients with muscle related symptoms<br>**Exclusion:**<br>Patients with non-statin related conditions which lead to elevated CK level 3 months prior to statin prescription<br>Patients prescribed nitrates and levothyroxine within 3 days after concurrent elevation of CK |
| NHS-UK Statin intolerance pathway<br>**(NHS-UK)** | EAS and NICE guidelines | Primary/ Secondary | **Inclusion:**<br>Patients with statins related muscle symptoms<br>Patient with CK level ≥ 4x the ULN (ULN for males is 250 IU/L and for females is 150 IU/L) and < 10x the ULN |

**Notes:** CRB: Combined rule-based; CK: Creatine kinase; ULN: Upper limit of normal; IU/L: International units per litre; NLA: National Lipid Association; EAS: European Atherosclerosis Society; SIMs: Statin induced myopathy; USA: United States of America; CCWG: Canadian Consensus Working Group; CK MB: Creatine kinase myocardial band; SAMT: Statin-associated muscle toxicity; NHS-UK: National Health Service-United Kingdom; NICE: National Institute for Health and Care Excellence

The prevalence of statin intolerance was calculated from the final reference standard classification. The demographic characteristics of patients in the reference standard was presented in a table. Area-based social advantage and disadvantage were classified using the Socio-Economic Indexes for Australia (SEIFA) 2021 Index of Relative Socio-Economic Advantage and Disadvantage (IRSAD), which is based on suburbs and localities (44).

### 2.5 Validation of the EHR-detectable electronic phenotypes of statin intolerance

In addition to the manual identification of patients with statin intolerance using the EAS definition, established rule-based electronic phenotyping algorithms were implemented to enable automated identification of statin intolerance within the ePBRN dataset. The algorithms were then validated against the manually identified reference standard by determining the true positives, false positives, true negatives, and false negatives. For the validation of each algorithm, the performance measures used were accuracy, sensitivity, specificity, positive predictive value (PPV), negative predictive value (NPV), balanced accuracy, F1 score and Matthews correlation coefficient (MCC). Accuracy is the proportion of patients correctly identified as statin tolerant or intolerant by each algorithm compared with the reference standard (the number of correct predictions over the total number of predictions) (45). Sensitivity is the proportion of patients with statin intolerance who were correctly identified by the algorithm (the number of true positives over the sum of the true positives and false negatives) (46). Specificity is the proportion of patients without statin intolerance who were correctly identified by the algorithm (the number of true negatives over the sum of true negatives and false positives) (46). The PPV is the proportion of patients identified as statin intolerant by the algorithm who were classified as statin intolerant by the reference standard (the number of true positives over the sum of the true positives and false positives) (46). The NPV is the proportion of patients identified as not statin intolerant by the algorithm who were classified as not statin intolerant by the reference standard (the number of true negatives over the sum of the true negatives and false negatives) (46). Balanced accuracy was calculated as the average of sensitivity and specificity to correctly identify statin tolerant and intolerant patients. The F1 score was calculated as the harmonic mean of sensitivity and PPV, while MCC was used to assess overall classification performance by incorporating true positives, true negatives, false positives, and false negatives. The reference standard, consisting of a subset of 1,369 patients, was considered the correct or true classification.

### 2.6 Ethical considerations

This study and the use of data from ePBRN was approved by the UNSW Human Research Ethics Advisory Panel (HC230066; 23 June 2023).

## 3. Results

### 3.1 Identification of EHR-detectable electronic phenotypes of statin intolerance

A comprehensive review identified EHR-detectable electronic phenotypes of statin intolerance. Six rule-based phenotyping algorithms were employed in this study, namely Minnesota Combined Rule-Based (CRB) algorithm (47), Japan statin induced myopathy (SIMs) (48), USA SIMs (16), Singapore SIMs (algorithms A, B, C, and D) (49), Japan statin-associated muscle toxicity (SAMT) (50), and NHS-UK statin intolerance pathway (51). An overview of the identified phenotyping algorithms is shown in **Table 1**.

### 3.2 Patient selection

The ePBRN dataset consisted of a total of 249,345 patients with 3,103,072 visits, of whom 18,377 patients with 110,147 visits were prescribed statins. After excluding patients who were younger than 18 years at the index date, there were a total of 18,371 patients with 110,083 visits. Following the application of further exclusion criteria 2,788 patients with 24,358 visits were excluded, leaving 15,583 general practice patients with 85,725 visits for analysis. Details of the patient selection process are shown in **Figure 3**.

**Figure 3:**
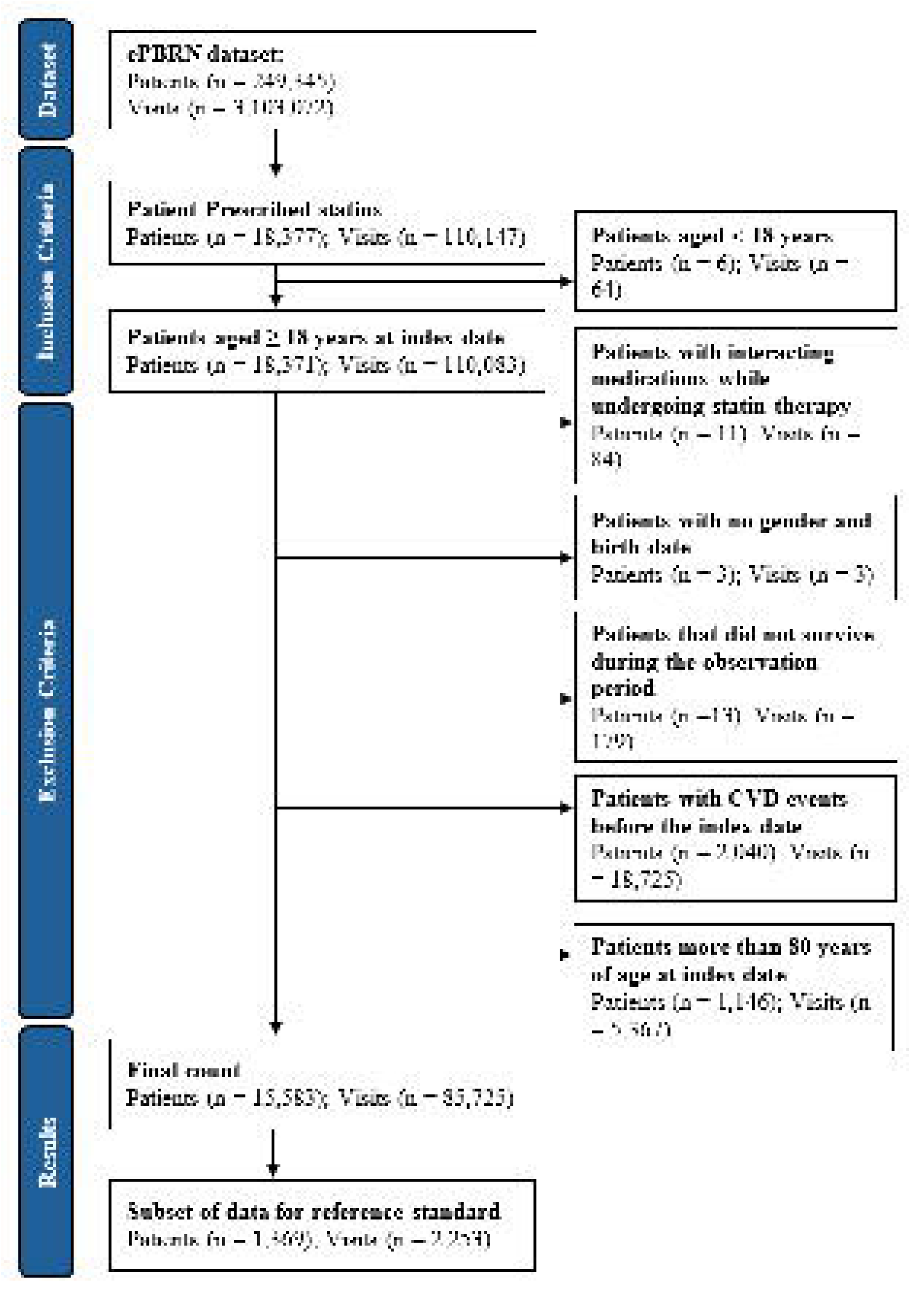
Identification of eligible patients from the ePBRN dataset. **Notes:** ePBRN: Electronic Practice Based Research Network; CVD: Cardiovascular disease

### 3.3 Manual case review and prevalence of statin intolerance

There were 1,369 patients in the reference standard cohort. Cohen’s kappa (IAA) between the two annotators was 0.894 **(Supplement 2)**. The DS for Annotator 1 was 0.968 and for Annotator 2 was 0.989, with an average DS of 0.978. The precision and recall for Annotator 1 were 0.944 and 0.994, and those for Annotator 2 were 0.985 and 0.992, respectively **(Supplement 3)**. Based on the manual case review using the reference standard, 794 patients were classified as statin intolerant, corresponding to a prevalence of 5.09% (794/15,583) among those prescribed statins in the ePBRN dataset.

### 3.4 Characteristics of patients with statin intolerance

The demographic characteristics of the patients in the reference standard is shown in **Table 2**. Statin intolerance was found in 794 patients. The mean age was 58.40 (±10.80) years for patients with statin intolerance and 55.80 (±12.40) years for the total patient population. The majority of the patients with statin intolerance were female (55.92%), aged 55–64 years (31.36%), non-smokers (50.50%) and residing in SEIFA IRSAD category 8 (34.63%). In comparison, among all patients, most were male (50.48%), within the same age group of 55–64 years (29.24%), non-smokers (41.96%) and predominantly from SEIFA IRSAD category 8 (34.76%).

**Table 2:** Demographic distribution of patients with statin intolerance.

| Variable | Category | Statin intolerant<br>(N = 794) | Total population<br>(N = 15,583) |
| --- | --- | --- | --- |
| **Age [mean (SD)]** |  | 58.40 (10.80) | 55.80 (12.40) |
| **Age**<br>**[n (%)]** | < 35 | 8 (1.01) | 815 (5.23) |
|  | 35 – 44 | 75 (9.45) | 2,208 (14.17) |
|  | 45 – 54 | 231 (29.09) | 4,171 (26.77) |
|  | 55 – 64 | 249 (31.36) | 4,556 (29.24) |
|  | 65 – 74 | 185 (23.30) | 2,920 (18.74) |
|  | ≥ 75 | 46 (5.79) | 913 (5.86) |
| **Gender**<br>**[n (%)]** | Male | 349 (43.95) | 7,866 (50.48) |
|  | Female or others | 445 (56.05) | 7,717 (49.52) |
| **Ethnicity**<br>**[n (%)]** | ATSI or not provided/recorded | 75 (9.45) | 4,309 (27.65) |
|  | Non-ATSI | 719 (90.55) | 11,274 (72.35) |
| **Employment**<br>**status [n (%)]** | Employed | 156 (19.65) | 3,471 (22.27) |
|  | Unemployed | 105 (13.22) | 2,577 (16.54) |
|  | Retired/pensioner | 160 (20.15) | 1,332 (8.55) |
|  | Not recorded | 373 (46.98) | 8,203 (52.64) |
| **SEIFA IRSAD**<br>**category**<br>**(decile)**<br>**[n (%)]** | 1 | 156 (19.65) | 1,703 (10.93) |
|  | 2 | 10 (1.26) | 1,046 (6.71) |
|  | 3 | 226 (28.46) | 3,772 (24.21) |
|  | 4 | 95 (11.96) | 2,688 (17.25) |
|  | 8 | 275 (34.63) | 5,417 (34.76) |
|  | 9 | 32 (4.03) | 957 (6.14) |
| **Smoking status**<br>**[n (%)]** | Smoker | 222 (27.96) | 4,112 (26.39) |
|  | Ex-smoker | 81 (10.20) | 1,076 (6.90) |
|  | Non-smoker | 401 (50.50) | 6,538 (41.96) |
|  | Not recorded | 90 (11.33) | 3,857 (24.75) |
| **Statin intensity at index date [n (%)] \*** | High | 164 (20.65) | 3,547 (22.76) |
|  | Moderate | 571 (71.91) | 10,507 (67.46) |
|  | Low | 51 (6.42) | 574 (3.68) |
| **Statin type at index date [n (%)] \*** | Atorvastatin | 308 (38.79) | 6,233 (40.00) |
|  | Rosuvastatin | 305 (38.41) | 5,941 (38.12) |
|  | Simvastatin | 132 (16.62) | 1,924 (12.35) |
|  | Fluvastatin or Pravastatin | 41 (5.16) | 530 (3.40) |

**Notes:**
SD: Standard deviation; ATSI: Aboriginals and Torres Strait Islanders; SEIFA: Socio-Economic Indexes for Australia; IRSAD: Index of Relative Socio-economic Advantage and Disadvantage (with lower categories indicating greater disadvantage and higher categories indicating greater advantage)
\* Variables with missing data. This includes 8 patients in statin intolerant group and 955 patients in total population group

### 3.5 Implementation and validation of the EHR-detectable electronic phenotyping algorithms for statin intolerance

The six rule-based phenotyping algorithms were Minnesota CRB algorithm, Japan SIMs, USA SIMs, Singapore SIMs (algorithms A, B, C, and D), Japan SAMT, and NHS-UK Statin intolerance pathway. The prevalence of statin intolerance, as identified by each of the algorithms is shown in **Figure 4**. The overlap in patients identified as statin intolerant by each phenotyping algorithm and the reference standard, as well as between the algorithms, is presented in **Supplement 4**.

**Figure 4:**
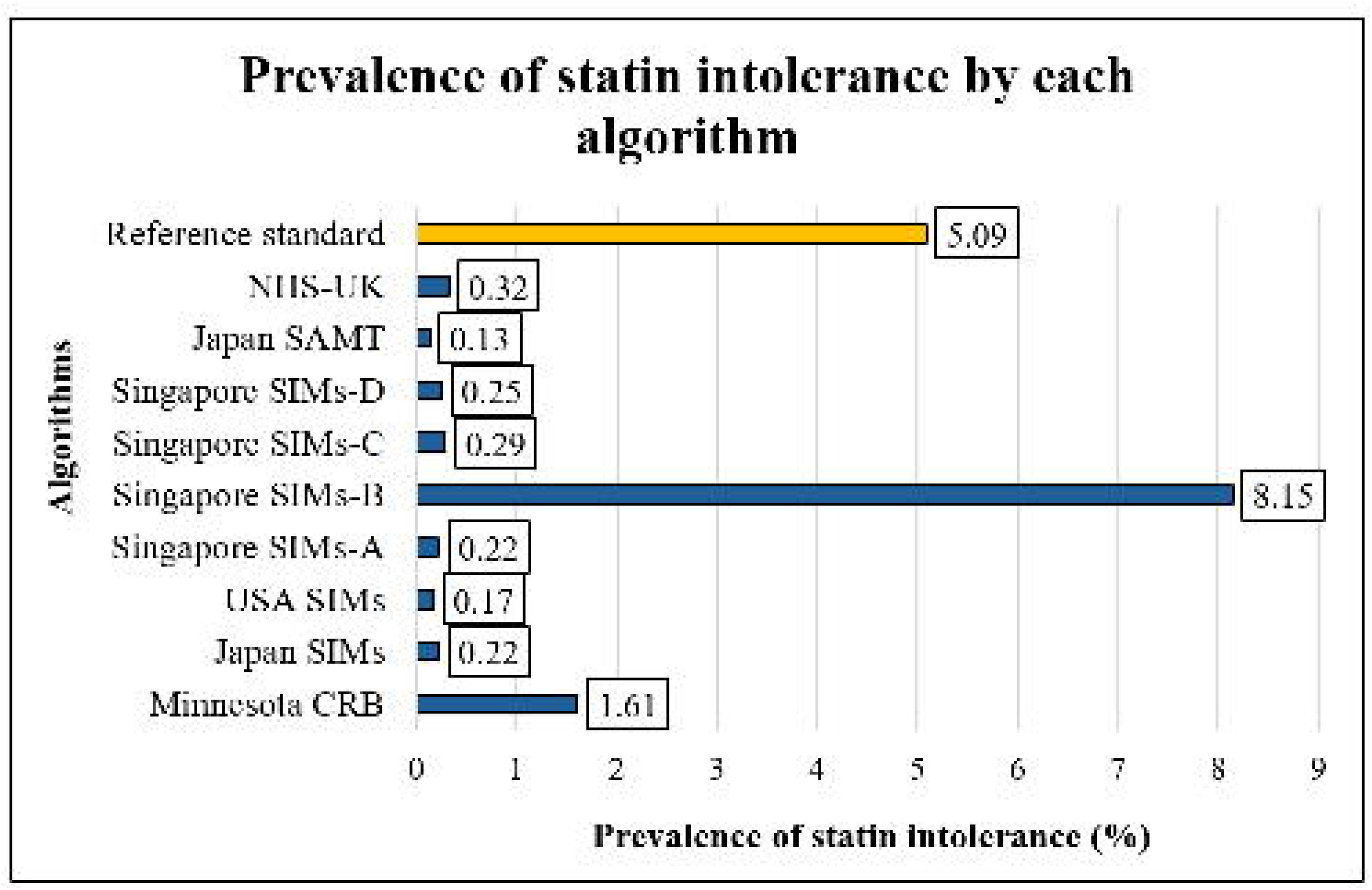
Prevalence of statin intolerance, as identified by each algorithm (N = 15,583) **Notes:** NHS-UK: National Health Service-United Kingdom; SAMT: Statin-associated muscle toxicity; SIMs: Statin induced myopathy; USA: United States of America; CRB: Combined rule-based

The validation results for each electronic phenotyping algorithm are shown in **Table 3**. Accuracy ranged from 42.07% to 57.05%, sensitivity from 1.76% to 92.95%, specificity from 7.48% to 99.13%, PPV from 51.85% to 76.19%, and NPV from 41.50% to 43.43%. The Singapore SIMs-B algorithm showed the highest accuracy, sensitivity, and NPV, while Japan SAMT showed the highest specificity and PPV. The true positives, true negatives, false positives and the false negatives for each algorithm are shown in **Supplement 5**.

**Table 3:**
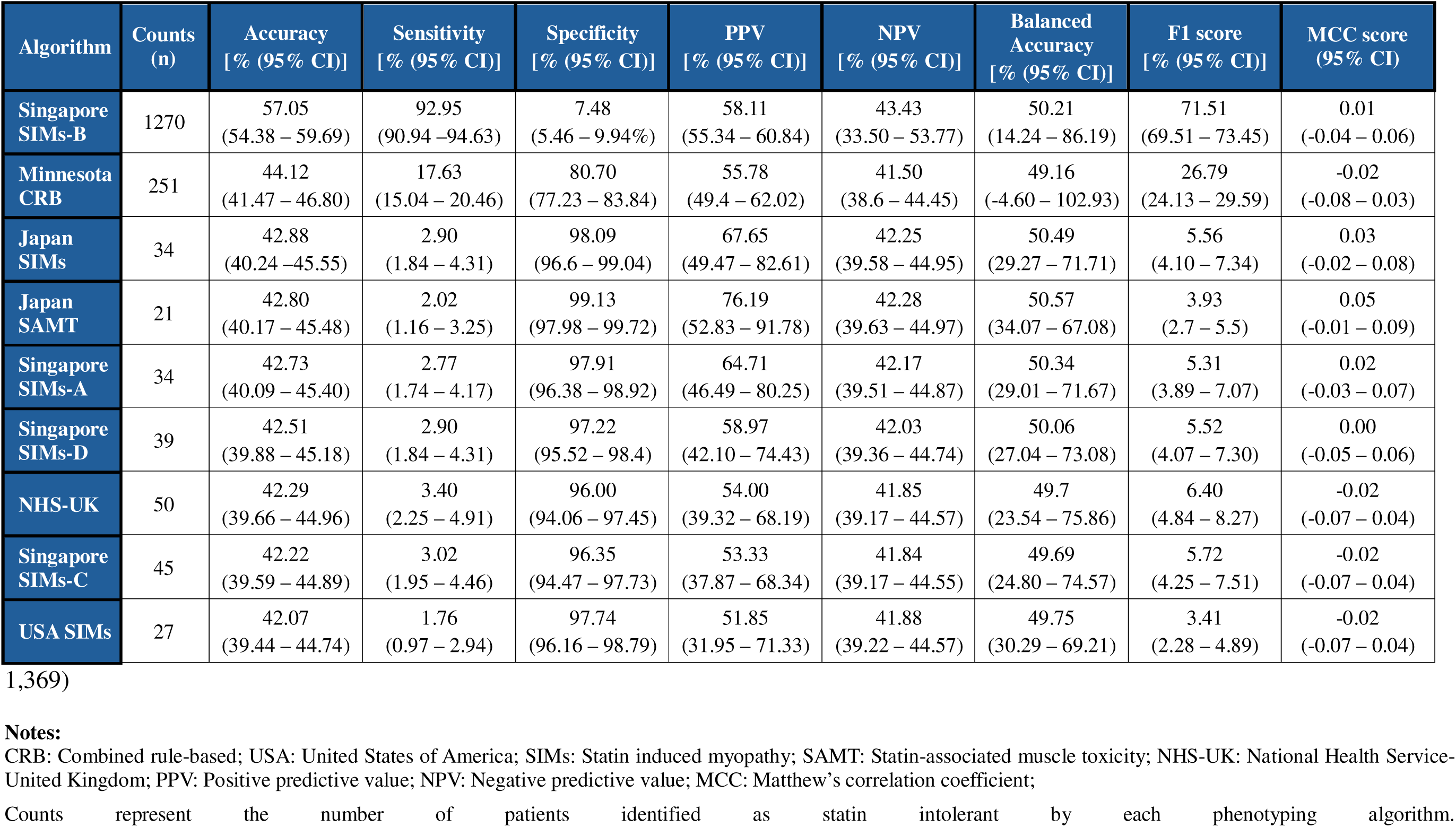
Summary performance metrics for each algorithm for identifying patients with statin intolerance compared to reference standard (N =.

| Algorithm | Counts<br>(n) | Accuracy<br>[% (95% CI)] | Sensitivity<br>[% (95% CI)] | Specificity<br>[% (95% CI)] | PPV<br>[% (95% CI)] | NPV<br>[% (95% CI)] | Balanced<br>Accuracy<br>[% (95% CI)] | F1 score<br>[% (95% CI)] | MCC score<br>(95% CI) |
| --- | --- | --- | --- | --- | --- | --- | --- | --- | --- |
| Singapore<br>SIMs-B | 1270 | 57.05<br>(54.38 – 59.69) | 92.95<br>(90.94 – 94.63) | 7.48<br>(5.46 – 9.94%) | 58.11<br>(55.34 – 60.84) | 43.43<br>(33.50 – 53.77) | 50.21<br>(14.24 – 86.19) | 71.51<br>(69.51 – 73.45) | 0.01<br>(-0.04 – 0.06) |
| Minnesota<br>CRB | 251 | 44.12<br>(41.47 – 46.80) | 17.63<br>(15.04 – 20.46) | 80.70<br>(77.23 – 83.84) | 55.78<br>(49.4 – 62.02) | 41.50<br>(38.6 – 44.45) | 49.16<br>(-4.60 – 102.93) | 26.79<br>(24.13 – 29.59) | -0.02<br>(-0.08 – 0.03) |
| Japan<br>SIMs | 34 | 42.88<br>(40.24 – 45.55) | 2.90<br>(1.84 – 4.31) | 98.09<br>(96.6 – 99.04) | 67.65<br>(49.47 – 82.61) | 42.25<br>(39.58 – 44.95) | 50.49<br>(29.27 – 71.71) | 5.56<br>(4.10 – 7.34) | 0.03<br>(-0.02 – 0.08) |
| Japan<br>SAMT | 21 | 42.80<br>(40.17 – 45.48) | 2.02<br>(1.16 – 3.25) | 99.13<br>(97.98 – 99.72) | 76.19<br>(52.83 – 91.78) | 42.28<br>(39.63 – 44.97) | 50.57<br>(34.07 – 67.08) | 3.93<br>(2.7 – 5.5) | 0.05<br>(-0.01 – 0.09) |
| Singapore<br>SIMs-A | 34 | 42.73<br>(40.09 – 45.40) | 2.77<br>(1.74 – 4.17) | 97.91<br>(96.38 – 98.92) | 64.71<br>(46.49 – 80.25) | 42.17<br>(39.51 – 44.87) | 50.34<br>(29.01 – 71.67) | 5.31<br>(3.89 – 7.07) | 0.02<br>(-0.03 – 0.07) |
| Singapore<br>SIMs-D | 39 | 42.51<br>(39.88 – 45.18) | 2.90<br>(1.84 – 4.31) | 97.22<br>(95.52 – 98.4) | 58.97<br>(42.10 – 74.43) | 42.03<br>(39.36 – 44.74) | 50.06<br>(27.04 – 73.08) | 5.52<br>(4.07 – 7.30) | 0.00<br>(-0.05 – 0.06) |
| NHS-UK | 50 | 42.29<br>(39.66 – 44.96) | 3.40<br>(2.25 – 4.91) | 96.00<br>(94.06 – 97.45) | 54.00<br>(39.32 – 68.19) | 41.85<br>(39.17 – 44.57) | 49.7<br>(23.54 – 75.86) | 6.40<br>(4.84 – 8.27) | -0.02<br>(-0.07 – 0.04) |
| Singapore<br>SIMs-C | 45 | 42.22<br>(39.59 – 44.89) | 3.02<br>(1.95 – 4.46) | 96.35<br>(94.47 – 97.73) | 53.33<br>(37.87 – 68.34) | 41.84<br>(39.17 – 44.55) | 49.69<br>(24.80 – 74.57) | 5.72<br>(4.25 – 7.51) | -0.02<br>(-0.07 – 0.04) |
| USA SIMs | 27 | 42.07<br>(39.44 – 44.74) | 1.76<br>(0.97 – 2.94) | 97.74<br>(96.16 – 98.79) | 51.85<br>(31.95 – 71.33) | 41.88<br>(39.22 – 44.57) | 49.75<br>(30.29 – 69.21) | 3.41<br>(2.28 – 4.89) | -0.02<br>(-0.07 – 0.04) |

CRB: Combined rule-based; USA: United States of America; SIMs: Statin induced myopathy; SAMT: Statin-associated muscle toxicity; NHS-UK: National Health Service- United Kingdom; PPV: Positive predictive value; NPV: Negative predictive value; MCC: Matthew's correlation coefficient;
Counts represent the number of patients identified as statin intolerant by each phenotyping algorithm.

Among the additional performance measures, the Singapore SIMs-B algorithm had the highest F1 score (71.51%). The Japan SAMT algorithm had the highest correlation with the reference standard as displayed by the MCC value of 0.046, although MCC values were close to zero across all algorithms, indicating weak overall correspondence with the reference standard.

The Japan SAMT algorithm had the highest precision–recall area under the curve (PR AUC) value (0.593) among the evaluated algorithms. The PR AUC value for each algorithm is shown in **Supplement 6**, and the precision–recall curves are presented in **Supplement 7**. A heatmap showing the performance metrics of each algorithm for identifying patients with statin intolerance compared to the reference standard is shown in **Supplement 8**.

## 4. Discussion

This study found that the prevalence of statin intolerance in the ePBRN dataset was 5.09%, which aligns marginally with the prevalence of statin intolerance in Australia which is between 5 and 15% of patients (7, 8). Prevalence estimates vary according to the definition, clinical setting, and method of case ascertainment, with lower estimates generally reported when more stringent definitions are applied (5, 52, 53, 54). Meta-analyses shows that the prevalence of statin intolerance can be as low as 3-5%, when strict definitions are used, and as high as 10-30% when broader definitions are employed (5, 53, 54). However, an additional consideration while interpreting the data is the nocebo/drucebo effect, which is a phenomenon where the adverse symptoms of statins are influenced by patient expectation and/or fear of intolerance (often influenced by negative representations by the media or close associates) (5, 55). This results in overestimation of intolerance, making it a crucial factor during the identification of patients with statin intolerance (56, 57, 58). In this study, cases in the reference standard were manually reviewed using the EAS definition based on information available in the EHR, including statin prescription records, CK values, and muscle symptoms. While CK provides an objective measure of muscle injury, the recording of muscle symptoms relies on patients reporting their symptoms to their GP. Therefore, muscle symptoms influenced by the nocebo/drucebo effect may have been recorded as potentially statin related symptoms, even when a causal relationship with statin therapy could not be established. Conversely, symptoms that were not reported by patients or documented by GPs would not have been captured in the EHR. These factors may have influenced the identification and classification of statin intolerance in the reference standard.

The six rule-based phenotyping algorithms used for the identification of patients with statin intolerance applied different combinations of clinical and EHR variables. Rule-based phenotyping provides a transparent approach to identifying clinical phenotypes from EHR data using predefined criteria, which may include diagnostic codes, medications, clinical observations, procedures, medical history, and laboratory results (30, 31, 59, 60, 61, 62). However, the performance of these algorithms depends on both the criteria used to define the phenotype and the availability and completeness of the required information within the EHR (30, 31, 63). In this study, none of the algorithms performed consistently well across both screening and confirmation domains.

The algorithms were developed across different clinical settings, which may affect their applicability to primary care EHR data. The Minnesota CRB algorithm was developed in primary care and relies on clinical events (such as treatment changes, discontinuations, and CK elevations), making its criteria relatively compatible with general practice EHR data (5). The Japan SIMs and Singapore SIMs algorithms were developed in hospital or tertiary care settings but use muscle symptoms, prescription information, and/or CK elevations that may also be captured in primary care EHRs (49, 53). The USA SIMs algorithm combines symptoms, adverse events, and therapy changes across different healthcare settings, allowing its criteria to be mapped to general practice data (54). In contrast, the Japan SAMT algorithm was primarily developed in the context of clinical trials and includes more detailed clinical inclusion and exclusion criteria that may be less consistently available in routinely collected general practice data (50). The NHS-UK algorithm is a management pathway rather than a patient identification algorithm but includes criteria that can be mapped to primary care EHR data where documented (64).

However, the availability and completeness of these variables may differ between primary and secondary or tertiary care settings. In primary care, CK measurements are only available when clinically requested, while muscle symptoms depend on their presentation and documentation in the EHR. In addition, treatment changes or care received outside participating ePBRN general practices may not be captured. These differences may have contributed to under-identification or misclassification of statin intolerance and variation in algorithm performance.

Algorithm performance varied according to the criteria used to define statin intolerance. Singapore SIMs-B, which relies primarily on muscle symptoms without requiring CK elevation, identified the most patients (n=1,270) and showed the highest accuracy (57.05%), sensitivity (92.95%), NPV (43.43%) and F1 (71.51%), but very low specificity (7.48%). This reflects a broad phenotype that may be more suitable for initial screening than confirmation (52, 65). In contrast, Minnesota CRB requires CK elevation greater than three times the ULN and showed lower sensitivity (17.63%) but higher specificity (80.70%), suggesting that its more restrictive CK-based criteria may miss patients with muscle symptoms without substantial CK elevation (5, 52).

The remaining algorithms identified relatively few patients (n=21 – 50) and generally showed very low sensitivity (1.76% – 3.40%) but high specificity (96.00% – 99.13%), consistent with their more restrictive criteria involving CK thresholds, temporal relationships, medication related factors, or exclusion of alternative causes (49, 50, 53, 54, 64). Japan SAMT, for example, had the highest specificity (99.13%) and PPV (76.19%) but sensitivity of only 2.02%. However, MCC values were close to zero across all algorithms (–0.02 to 0.05), indicating weak overall correspondence with the reference standard. Therefore, the high sensitivity of Singapore SIMs-B and high specificity of Japan SAMT should be interpreted as strengths for different classification purposes rather than evidence of strong overall performance. A two-stage approach using a sensitive algorithm for initial screening followed by a more specific algorithm for further assessment may warrant further investigation, although additional validation is required before clinical implementation (66, 67, 68).

Overall, the findings demonstrate that the identification of patients with statin intolerance is influenced by the criteria used to define intolerance and the availability of the required information within EHR data. Accurate identification of statin intolerance is important because intolerance may contribute to statin discontinuation or poor statin adherence, potentially affecting the effectiveness of cardiovascular prevention (69). The findings suggest that a combination of algorithms with complementary performance characteristics may warrant further investigation. In particular, the Singapore SIMs-B algorithm may have potential as an initial screening approach because of its high sensitivity, while the higher specificity of the Japan SAMT algorithm may provide complementary information during further assessment. However, the low MCC values across all algorithms indicate that none demonstrated strong overall classification performance. Further validation and refinement are therefore required before these algorithms, individually or in combination, can be recommended for clinical application.

### Limitations

1. **The reference standard was developed from a preselected subset of patients rather than the entire study population.** Manual review using the EAS definition was conducted among 1,369 patients who had been identified as potential cases of statin intolerance by at least one of the electronic phenotyping algorithms. Consequently, patents with intolerance who were not identified by any of the algorithms would not have been included in the reference standard assessment. Therefore, the reference standard subset may not have captured all cases of statin intolerance in the eligible population, which could have potentially underestimated the prevalence and affected estimates of algorithm performance, particularly specificity and NPV.
2. **There is no universally accepted reference standard for identifying statin intolerance using EHR data.** The reference standard developed through manual review and application of the EAS definition provided a clinically informed reference but should not be considered a definitive gold standard. Classification depended on routinely recorded EHR information, and incomplete documentation of symptoms, laboratory results, and treatment changes may have resulted in misclassification.
3. **Definitions and criteria for statin intolerance varied across the electronic phenotyping algorithms.** The algorithms differed in their use of muscle symptoms, CK thresholds, treatment changes, and exclusion of alternative causes, which may have contributed to variation in algorithm performance and estimated prevalence. Although sex-specific CK thresholds were considered, variation in CK levels according to ethnicity was not incorporated and may also have resulted in misclassification.
4. **The availability and completeness of clinical information in primary care EHRs may have affected algorithm performance.** Several algorithms were originally developed using secondary or tertiary care data, where clinical and laboratory information may be more systematically available. Incomplete recording of CK measurements, muscle symptoms, medication changes, or care received outside the participating ePBRN general practices may have resulted in under-identification or misclassification. Moreover, the index date reflected the first statin prescription recorded in ePBRN and may not represent treatment initiation if earlier prescriptions occurred outside participating practices. Missing data for some patient characteristics also limited the completeness with which the study population could be described.
5. **Recorded statin prescriptions do not confirm that patients obtained or took the prescribed medication.** As linked dispensing data were unavailable, patients with limited or no exposure to statins may have been included despite having no opportunity to experience statin related adverse effects, potentially contributing to underestimation of statin intolerance.
6. **The exclusion criteria may have introduced selection bias.** Patients who experienced a CVD event during follow-up were excluded to maintain a primary prevention cohort, although they were receiving statins for primary prevention before the event. This exclusion may have affected the representativeness of the cohort.
7. **The findings may have limited generalizability beyond the study population and setting.** The study included patients receiving statins for primary prevention using routinely collected EHR data from general practices in SWS. The findings may therefore not be directly applicable to secondary prevention populations, other healthcare systems, or populations with different demographic characteristics, prescribing practices, and EHR structures. Further validation in other populations and settings is required.

## 5. Conclusion

This study found that the prevalence of statin intolerance in the ePBRN dataset was 5.09%. None of the evaluated rule-based electronic phenotyping algorithms performed consistently well across both screening and confirmation domains. The Singapore SIMs-B algorithm displayed the highest accuracy (57.05%), sensitivity (92.95%), NPV (43.43%) and F1 (71.51%) scores, while the Japan SAMT algorithm displayed the highest specificity (99.13%), PPV (76.19%) and MCC (0.05). Based on these metrics, it can be inferred that in practice it might be logical to use a two-stage approach involving screening with Singapore SIMs-B to flag patient with potential statin intolerance, followed by assessment using Japan SAMT as a confirmation step, before making definitive management changes. This would align with a diagnostic workflow that emphasizes minimizing missed cases and avoiding unnecessary treatment changes. The selection of an algorithm, or a combination of algorithms, for the identification of patients with statin intolerance depends significantly on several factors, and understanding these factors may be the key to identifying these patients. The results of this study emphasise that EHR-detectable phenotypes should be used as decision support aids rather than as definitive diagnostic tools and that clinical judgement and patient engagement are necessary for the management of suspected statin intolerance. Overall, the findings suggest that the performance and potential utility of electronic phenotyping algorithms depend on their underlying criteria and the context in which they are applied. Further validation is required before these algorithms can be used to support the identification of statin intolerance in clinical practice.

## Author contributions

1. SR and JJ contributed to the conceptualization of this study.
2. JJ contributed to the funding acquisition
3. JR, KAR, STL and JJ contributed to the resources and supervision of this study
4. JR, KAR, STL and JJ contributed to the validation
5. All authors contributed to the reviewing and editing of this study
6. SR and JJ contributed to data curation
7. SR contributed to the formal analysis and the original drafting of this study
8. SR contributed to the methodology of the study

## Data sharing statement

The corresponding author will make the data available upon request.

## Data Availability

The corresponding author will make the data available up on request.

## Acknowledgements

We would like to thank the ePBRN Primary Care Health Informatics Working Group of the Secure Research Environment for Digital Health (SREDH) Consortium (www.sredhconsortium.org, accessed on 7 October 2024) for their assistance with access to the ePBRN dataset to investigate the findings from this review.

