## Supplement for "Electronic health record (EHR)-detectable phenotypes of statin intolerance: Prevalence and validation in real-world general practice"

**Supplement 1:** The RECORD statement – checklist of items, extended from the STROBE statement, that should be reported in observational studies using routinely collected health data.

|  | **Item No.** | **STROBE items** | **Location in manuscript where items are reported** | **RECORD items** | **Location in manuscript where items are reported** |
| --- | --- | --- | --- | --- | --- |
| **Title and abstract** | | |  |  |  |
|  | 1 | (a) Indicate the study’s design with a commonly used term in the title or the abstract (b) Provide in the abstract an informative and balanced summary of what was done and what was found | (a) Page 2  (b) Page 2 | RECORD 1.1: The type of data used should be specified in the title or abstract. When possible, the name of the databases used should be included.  RECORD 1.2: If applicable, the geographic region and timeframe within which the study took place should be reported in the title or abstract.  RECORD 1.3: If linkage between databases was conducted for the study, this should be clearly stated in the title or abstract. | 1.1 Page 2  1.2 Not applicable  1.3 Not applicable |
| **Introduction** | | |  |  |  |
| Background rationale | 2 | Explain the scientific background and rationale for the investigation being reported | Pages 3-5 |  |  |
| Objectives | 3 | State specific objectives, including any prespecified hypotheses | Page 5 |  |  |
| **Methods** | | |  |  |  |
| Study Design | 4 | Present key elements of study design early in the paper | Page 8-9 |  |  |
| Setting | 5 | Describe the setting, locations, and relevant dates, including periods of recruitment, exposure, follow-up, and data collection | Pages 7-8 |  |  |
| Participants | 6 | *(a) Cohort study* - Give the  eligibility criteria, and the sources and methods of selection of participants. Describe methods of follow-up  *Case-control study* - Give the eligibility criteria, and the sources and methods of case ascertainment and control selection. Give the rationale for the choice of cases and controls *Cross-sectional study* - Give the eligibility criteria, and the sources and methods of selection of participants  *(b) Cohort study* - For matched studies, give matching criteria and number of exposed and unexposed  *Case-control study* - For matched studies, give matching criteria and the number of controls per case | (a) Pages 7-8  (b) Not applicable | RECORD 6.1: The methods of study  population selection (such as codes or algorithms used to identify subjects) should be listed in detail. If this is not possible, an explanation should be provided.  RECORD 6.2: Any validation studies of the codes or algorithms used to select the population should be referenced. If validation was conducted for this study and not published elsewhere, detailed methods and results should be provided.  RECORD 6.3: If the study involved linkage of databases, consider use of a flow diagram or other graphical display to demonstrate the data linkage process, including the number of individuals with linked data at each stage. | 6.1 Pages 7-8  6.2 Not applicable  6.3 Page 6 |
| Variables | 7 | Clearly define all outcomes, exposures, predictors, potential confounders, and effect modifiers. Give diagnostic criteria, if applicable. | Pages 10-12 | RECORD 7.1: A complete list of codes and algorithms used to classify exposures, outcomes, confounders, and effect modifiers should be provided. If these cannot be reported, an explanation should be provided. | Pages 10-12 |
| Data sources/ measurement | 8 | For each variable of interest, give sources of data and details of methods of assessment (measurement).  Describe comparability of assessment methods if there is more than one group | Pages 7-8 |  |  |
| Bias | 9 | Describe any efforts to address potential sources of bias | Not applicable |  |  |
| Study size | 10 | Explain how the study size was arrived at | Not applicable |  |  |
| Quantitative variables | 11 | Explain how quantitative variables were handled in the analyses. If applicable, describe which groupings were chosen, and why | Pages 7-8 and pages 10-11 |  |  |
| Statistical  methods | 12 | (a) Describe all statistical methods, including those used to control for confounding (b) Describe any methods used to examine subgroups and interactions   (c) Explain how missing data were addressed (d) *Cohort study* - If applicable, explain how loss to follow-up   was addressed  *Case-control study* - If applicable, explain how matching of cases and controls  was addressed  *Cross-sectional study* - If applicable, describe analytical methods taking account of sampling strategy   (e) Describe any sensitivity analyses | (a) Not applicable  (b) Not applicable  (c) Not applicable  (d) Pages 9-11  (e) Not applicable |  |  |
| Data access and cleaning methods |  | .. |  | RECORD 12.1: Authors should describe the extent to which the investigators had access to the database population used to create the study population.  RECORD 12.2: Authors should provide information on the data cleaning methods used in the study. | 12.1 Page 7-8  12.2 Not applicable |
| Linkage |  | .. |  | RECORD 12.3: State whether the | Page 7 |
|  |  |  |  | study included person-level, institutional-level, or other data linkage across two or more databases. The methods of linkage and methods of linkage quality evaluation should be provided. |  |
| **Results** | | | | | |
| Participants | 13 | (a) Report the numbers of individuals at each stage of the study (*e.g.*, numbers potentially eligible, examined for eligibility, confirmed eligible, included in the study, completing follow-up, and analysed) (b) Give reasons for nonparticipation at each stage. (c) Consider use of a flow diagram | (a) Pages 17-18  (b) Not applicable  (c) Page 18 | RECORD 13.1: Describe in detail the selection of the persons included in the study (*i.e.,* study population selection) including filtering based on data quality, data availability and linkage. The selection of included persons can be described in the text and/or by means of the study flow diagram. | Pages 17-18 |
| Descriptive data | 14 | (a) Give characteristics of study participants (*e.g.*, demographic, clinical, social) and information on exposures and potential confounders (b) Indicate the number of participants with missing data for each variable of interest (c) *Cohort study* - summarise follow-up time (*e.g.*, average and total amount) | (a) Pages 19-20  (b) Pages 19-20  (c) Not applicable |  |  |
| Outcome data | 15 | *Cohort study* - Report numbers of outcome events or summary measures over time  *Case-control study* - Report numbers in each exposure category, or summary measures of exposure  *Cross-sectional study* - Report numbers of outcome events or  summary measures | Pages 19-20 |  |  |
| Main results | 16 | (a) Give unadjusted estimates and, if applicable, confounder adjusted estimates and their precision (e.g., 95% confidence interval). Make clear which confounders were adjusted for and why they were included (b) Report category boundaries when continuous variables were categorized  (c) If relevant, consider translating estimates of relative risk into absolute risk for a meaningful time period | (a) Not applicable  (b) Pages 19-20  (c) Not applicable |  |  |
| Other analyses | 17 | Report other analyses done—e.g., analyses of subgroups and interactions, and sensitivity analyses | Pages 21-23 |  |  |
| **Discussion** | | | | | |
| Key results | 18 | Summarise key results with reference to study objectives | Page 24 |  |  |
| Limitations | 19 | Discuss limitations of the study, taking into account sources of potential bias or imprecision. Discuss both direction and magnitude of any potential bias | Pages 26-28 | RECORD 19.1: Discuss the  implications of using data that were not created or collected to answer the specific research question(s). Include discussion of misclassification bias, unmeasured confounding, missing data, and changing eligibility over time, as they pertain to the study being reported. | Pages 26-28 |
| Interpretation | 20 | Give a cautious overall interpretation of results considering objectives, limitations, multiplicity of analyses, results from similar studies, and other relevant evidence | Pages 24-26 |  |  |
| Generalisability | 21 | Discuss the generalisability (external validity) of the study results | Page 28 |  |  |
| **Other Information** | | | | | |
| Funding | 22 | Give the source of funding and the role of the funders for the present study and, if applicable, for the original study on which the present article is based | Page 30 |  |  |
| Accessibility of protocol, raw data, and programming code |  |  | Page 30 | RECORD 22.1: Authors should provide information on how to access any supplemental information such as the study protocol, raw data, or programming code. | Supplement 1-8 |

*Reference: Benchimol EI, Smeeth L, Guttmann A, Harron K, Moher D, Petersen I, Sørensen HT, von Elm E, Langan SM, the RECORD Working Committee. The REporting of studies Conducted using Observational Routinely-collected health Data (RECORD) Statement. *PLoS Medicine* 2015; in press.

*Checklist is protected under Creative Commons Attribution ([CC BY](http://creativecommons.org/licenses/by/4.0/)) license.

**Supplement 2:** The calculated Inter Annotation Agreement (IAA) of the two researchers (annotators) for the reference standard

|  | **Agreements** | **Conflicts** | **Kappa** | **z** | **p-value** |
| --- | --- | --- | --- | --- | --- |
| **IAA** | 1296 | 73 | 0.894 | 33.1 | 0.00 |

**Supplement 3:** The calculated Deviation Score (DS) of the two researchers (annotators) for the reference standard

|  | **Annotator 1** | **Annotator 2** |
| --- | --- | --- |
| **Precision** | 0.944 | 0.985 |
| **Recall** | 0.994 | 0.992 |
| **DS** | 0.968 | 0.989 |
| **Average DS** | 0.978 | |

**Supplement 4:** Table showing overlap in patients identified as statin intolerant by the phenotyping algorithms and the reference standard

| **Algorithm** | **Reference standard** | **Minnesota CRB** | **USA SIMs** | **Japan SIMs** | **Singapore SIMs-A** | **Singapore SIMs-B** | **Singapore SIMs-C** | **Singapore SIMs-D** | **Japan SAMT** | **NHS-UK** |
| --- | --- | --- | --- | --- | --- | --- | --- | --- | --- | --- |
| **Reference standard** | 794 | 140 | 14 | 23 | 22 | 738 | 24 | 23 | 16 | **27** |
| **Minnesota CRB** | 140 | 251 | 0 | 0 | 0 | 231 | 10 | 10 | 3 | 15 |
| **USA SIMs** | 14 | 0 | 27 | 0 | 0 | 25 | 1 | 1 | 0 | 0 |
| **Japan SIMs** | 23 | 0 | 0 | 34 | 0 | 33 | 0 | 0 | 2 | 0 |
| **Singapore SIMs-A** | 22 | 0 | 0 | 0 | 34 | 30 | 2 | 2 | 0 | 2 |
| **Singapore SIMs-B** | 738 | 231 | 25 | 33 | 30 | 1270 | 0 | 0 | 19 | 47 |
| **Singapore SIMs-C** | 24 | 10 | 1 | 0 | 2 | 0 | 45 | 0 | 1 | 1 |
| **Singapore SIMs-D** | 23 | 10 | 1 | 0 | 2 | 0 | 0 | 39 | 0 | 2 |
| **Japan SAMT** | 16 | 3 | 0 | 2 | 0 | 19 | 1 | 0 | 21 | 0 |
| **NHS-UK** | 27 | 15 | 0 | 0 | 2 | 47 | 1 | 2 | 0 | 50 |

CRB: Combined rule-based; USA: United States of America; SIMs: Statin induced myopathy; SAMT: Statin-associated muscle toxicity; NHS-UK: National Health Service-United Kingdom.

**Supplement 5:** Table showing true positives, true negatives, false positives and false negatives of each algorithm for identifying patients with statin intolerance compared to the reference standard

| **Algorithm** | **Count (n)** | **True Positives** | **True Negatives** | **False Positives** | **False Negatives** |
| --- | --- | --- | --- | --- | --- |
| **Minnesota CRB** | 251 | 140 | 464 | 111 | 654 |
| **USA SIMs** | 27 | 14 | 562 | 13 | 780 |
| **Japan SIMs** | 34 | 23 | 564 | 11 | 771 |
| **Singapore SIMs-A** | 34 | 22 | 563 | 12 | 772 |
| **Singapore SIMs-B** | 1270 | 738 | 43 | 532 | 56 |
| **Singapore SIMs-C** | 45 | 24 | 554 | 21 | 770 |
| **Singapore SIMs-D** | 39 | 23 | 559 | 16 | 771 |
| **Japan SAMT** | 21 | 16 | 570 | 5 | 778 |
| **NHS-UK** | 50 | 27 | 552 | 23 | 767 |

CRB: Combined rule-based; USA: United States of America; SIMs: Statin induced myopathy; SAMT: Statin-associated muscle toxicity; NHS-UK: National Health Service-United Kingdom.

| **Algorithm** | **t** | **p-value** | **kappa** | **PR AUC**  **(95% CI)** |
| --- | --- | --- | --- | --- |
| **Minnesota CRB** | -0.789 | 0.430 | -0.0148 | 0.587  (0.56 - 0.61) |
| **USA SIMs** | -0.653 | 0.514 | -0.0042 | 0.583  (0.56 - 0.61) |
| **Japan SIMs** | 1.154 | 0.249 | 0.00832 | 0.591  (0.56 - 0.62) |
| **Singapore SIMs-A** | 0.802 | 0.423 | 0.00578 | 0.588  (0.56 - 0.62) |
| **Singapore SIMs-B** | 0.300 | 0.765 | 0.0048 | 0.585  (0.56 - 0.61) |
| **Singapore SIMs-C** | -0.644 | 0.519 | -0.0053 | 0.583  (0.56 - 0.61) |
| **Singapore SIMs-D** | 0.125 | 0.900 | 0.0010 | 0.585  (0.56 - 0.61) |
| **Japan SAMT** | 1.703 | 0.089 | 0.0097 | 0.593  (0.56 - 0.62) |
| **NHS-UK** | -0.583 | 0.560 | -0.0051 | 0.583  (0.56 - 0.61) |

**Supplement 6:** Table showing area under the curve values of each algorithm for identifying patients with statin intolerance compared to the reference standard

CRB: Combined rule-based; USA: United States of America; SIMs: Statin induced myopathy; SAMT: Statin-associated muscle toxicity; NHS-UK: National Health Service-United Kingdom; PR: Precision–recall; AUC: Area under the curve

**Supplement 7:** The Area under the curve (AUC) for each algorithm as per Precision–recall (PR) AUC

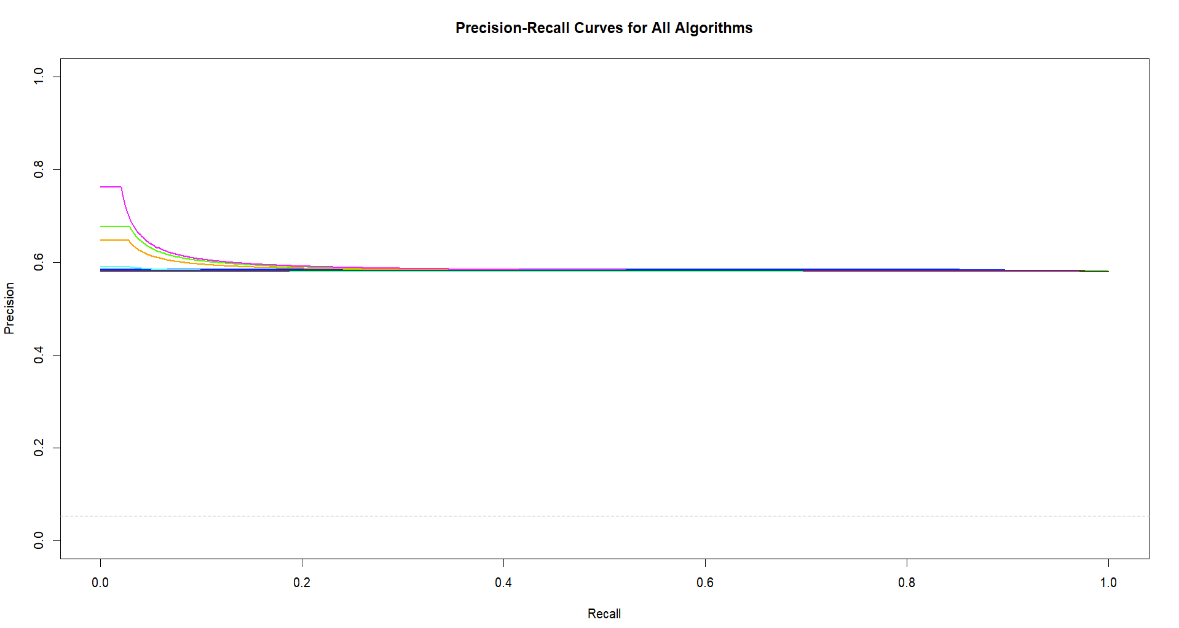

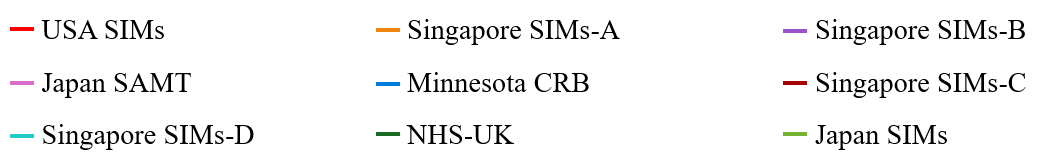

USA: United States of America; SIMs: Statin induced myopathy; SAMT: Statin-associated muscle toxicity; CRB: Combined rule-based; NHS-UK: National Health Service-United Kingdom.

**Supplement 8:** Heatmap table showing the performance metrics of each algorithm for identifying patients with statin intolerance compared to the reference standard

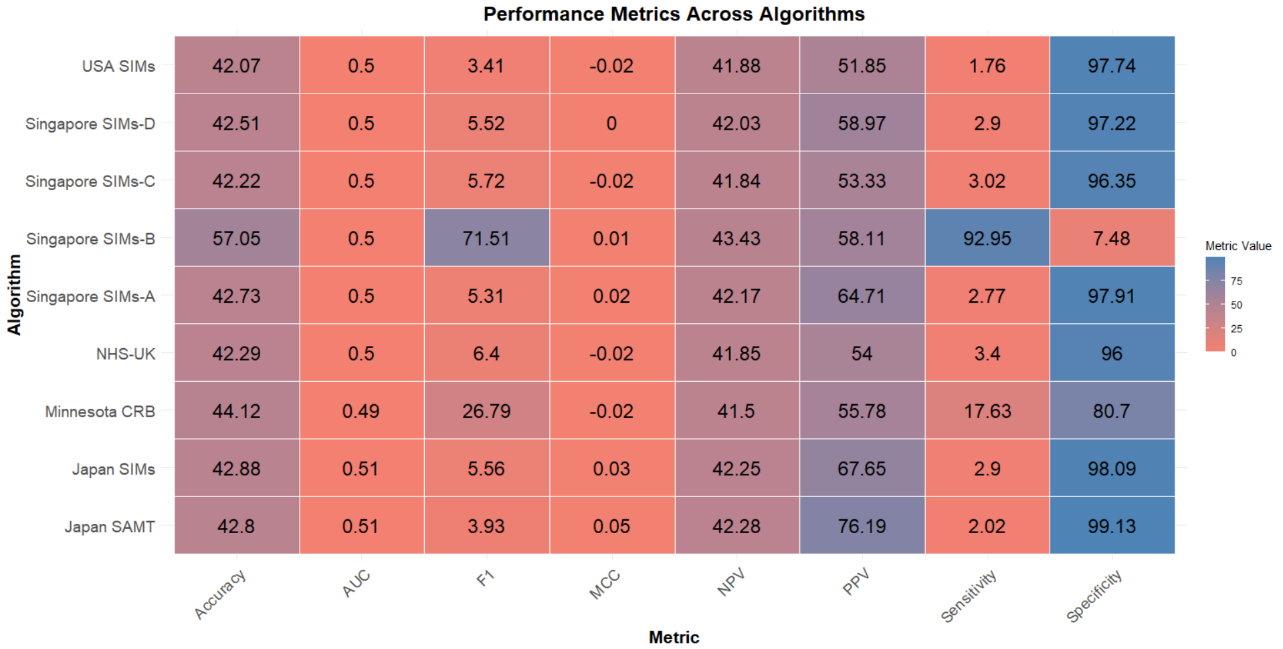

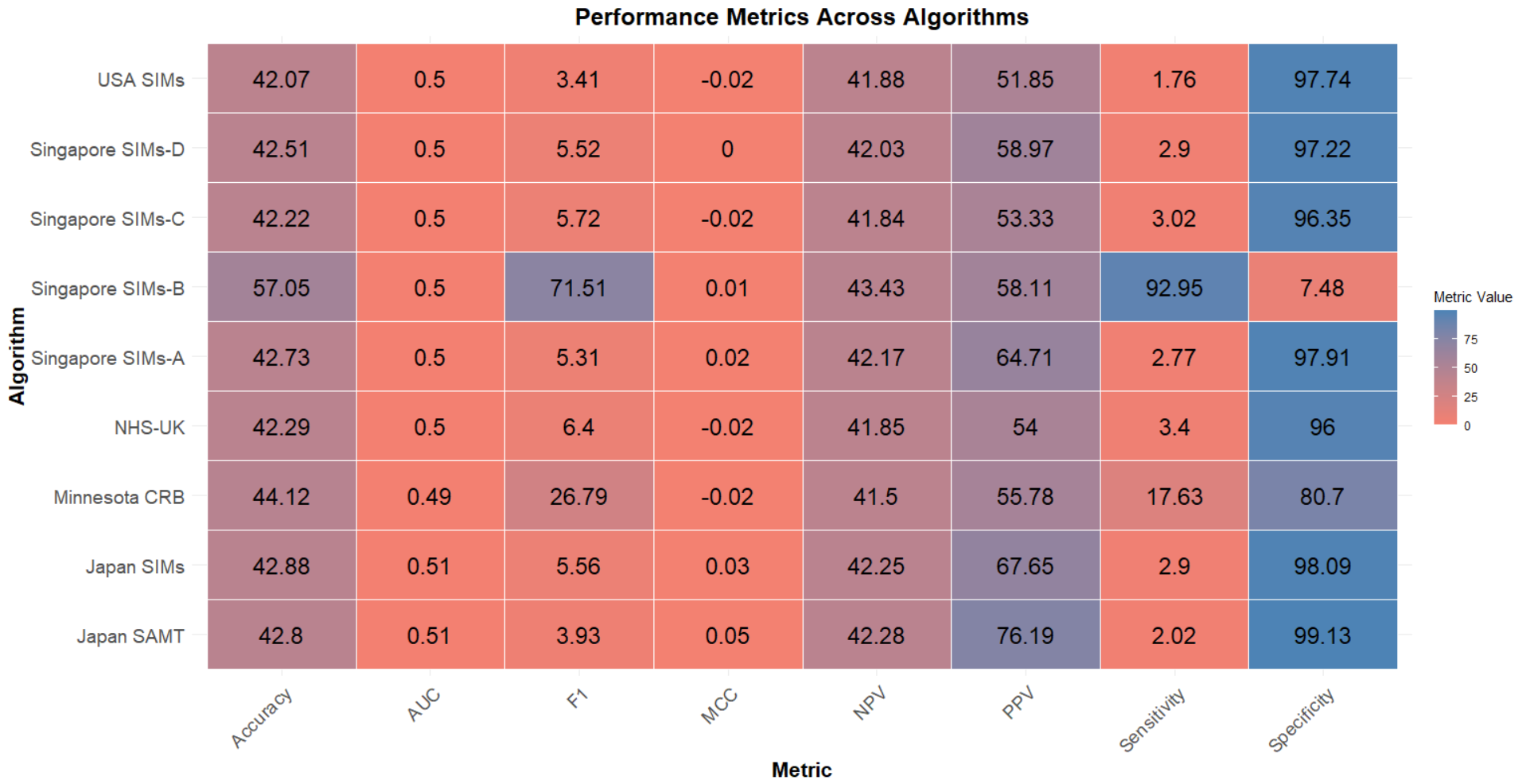
